# Genomic and mobility data reveal mass population movement as a driver of SARS-CoV-2 dissemination and diversity in Bangladesh

**DOI:** 10.1101/2021.01.05.21249196

**Authors:** Lauren A. Cowley, Mokibul Hassan Afrad, Sadia Isfat Ara Rahman, Md. Mahfuz-Al-Mamun, Taylor Chin, Ayesha Mahmud, Mohammed Ziaur Rahman, Mallick Masum Billah, Manjur Hossain Khan, Sharmin Sultana, Tilovatul Khondaker, Stephen Baker, Nandita Banik, Ahmed Nawsher Alam, Kaiissar Mannor, Sayera Banu, Anir Chowdhury, Meerjady Sabrina Flora, Nicholas R. Thomson, Caroline O. Buckee, Firdausi Qadri, Tahmina Shirin

## Abstract

**Background:** New data streams are being used to track the pandemic of SARS-CoV-2, including genomic data which provides insights into patterns of importation and spatial spread of the virus, as well as population mobility data obtained from mobile phones. Here, we analyse the emergence and outbreak trajectory of SARS-CoV-2 in Bangladesh using these new data streams, and identify mass population movements as a key early event driving the ongoing epidemic.

**Methods:** We sequenced complete genomes of 67 SARS-CoV-2 samples (March-July 2020) and combined this dataset with 324 genomes from Bangladesh. For phylogenetic context, we also used 68,000 GISAID genomes collected globally. We paired this genomic data with population mobility information from Facebook and three mobile phone operators.

**Findings:** The majority (85%) of the Bangladeshi sequenced isolates fall into either pangolin lineage B.1.36 (8%), B.1.1 (19%) or B.1.1.25 (58%). Bayesian time-scaled phylogenetic analysis predicted SARS-COV-2 first appeared in mid-February, through international introductions. The first case was reported on March 8^th^. This pattern of repeated international introduction changed at the end of March when three discrete lineages expanded and spread clonally across Bangladesh. The shifting pattern of viral diversity across Bangladesh is reflected in the mobility data which shows the mass migration of people from cities to rural areas at the end of March, followed by frequent travel between Dhaka and the rest of the country during the following months.

**Interpretation:** In Bangladesh, population mobility out of Dhaka as well as frequent travel from urban hotspots to rural areas resulted in rapid country-wide dissemination of SARS-CoV-2. The strains in Bangladesh reflect the local expansion of global lineages introduced early from international travellers to and from major international travel hubs. Importantly, the Bangladeshi context is consistent with epidemiologic and phylogenetic findings globally. Bangladesh is one of the few countries in the world with a rich history of conducting mass vaccination campaigns under complex circumstances. Combining genomics and these new data streams should allow population movements to be modelled and anticipated rendering Bangladesh extremely well prepared to immunize citizens rapidly. Based on our genomics data and the country’s successful immunization history, vaccines becoming available globally will be suitable for implementation in Bangladesh while ongoing genomic surveillance is conducted to monitor for new variants of the virus.

**Funding:** Government of Bangladesh, Bill and Melinda Gates Foundation, Wellcome Trust.

**Research in context:** *Evidence before this study:* The emergence of SARS-CoV-2, leading to the COVID-19 pandemic, has motivated all countries in the world to obtain high resolution data on the virus. Globally over 300,000 strains have been sequenced and information made available in GISAID. Within the first 100 days of the emergence of SARS-CoV-2, genomic analysis from different countries led to the development of vaccines which have now reached market. Information on the prevailing genotypes of SARS-CoV-2 since introduction is needed in low and middle-income countries (LMICs), including Bangladesh, in order to determine the suitability of therapeutics and vaccines in the pipeline and help vaccine deployment.

*Added value of this study:* We sequenced SARS-CoV-2 genomes from strains that were prospectively collected during the height of the pandemic and combined these genomic data with mobility data to comprehensively describe i) how repeated international importations of SARS-CoV-2 were ultimately linked to nationwide spread, ii) 85% of strains belonged to the Pangolin lineages B.1.1, B.1.1.25 and B.1.36 and that similar mutation rates were observed as seen globally iii) the switch in genomic dynamics of SARS-CoV-2 coincided with mass migration out of cities to the rest of the country. We have assessed the contributions of population mobility on the maintenance and spread of clonal lineages of SARS-CoV-2. This is the first time these data types have been combined to look at the spread of this virus nationally.

*Implications of all the available evidence:* SARS-CoV-2 genomic diversity and mutation rate in Bangladesh is comparable to strains circulating globally. Notably, the data on the genomic changes of SARS-CoV-2 in Bangladesh is reassuring, suggesting that immunotherapeutic and vaccines being developed globally should also be suitable for this population. Since Bangladesh already has extensive experience of conducting mass vaccination campaigns, such as the rollout of the oral Cholera vaccine, experience of developing and using new data streams will enable efficient and targeted immunization of the population in 2021 with COVID-19 vaccine(s).

## Introduction

The rapid global dissemination of SARS-CoV-2 from its origin in Wuhan, China, in late 2019 has now led to >85 million cases and >1·8 million deaths within twelve months. Initially, spread was strongly linked to international air travel with outbreaks in South–East Asia, Europe, and North America dominating global case counts in late spring of 2020. In an unprecedented global response, many countries, including Bangladesh, acted decisively and rapidly to restrict population movement and introduce additional social and behavioural interventions, all designed to slow the spread of the virus. Until now the impact of these policies has been hard to assess, in part because of the near-universal difficulties that countries have had rapidly scaling up RT-PCR testing capacity.

Bangladesh is a low and middle-income country (LMIC) with a population of over 166 million people, of which 63% live in rural regions.^1,2^ The first confirmed case of SARS-CoV-2 in Bangladesh was reported on March 8^th^ by the Institute of Epidemiology Disease Control and Research (IEDCR). To reduce community transmission, the Government of Bangladesh announced an official National General Holiday on March 23^rd^, effective from March 26^th^ to April 4^th^, then extended gradually until May 30^th^. As of December 21^st^, there were >500,000 confirmed COVID-19 cases.^3^ Whilst testing capacity was rapidly expanded, training and infrastructure for accurate epidemiological surveillance, particularly outside the capital Dhaka, remained challenging. As a result, it has been unclear how the epidemic spread in Bangladesh, how many regions remain susceptible, and what this means for the future trajectory of SARS-CoV-2 and the deployment of interventions, including therapeutics and vaccines.

Viral genomics approaches have been used to track the epidemic, enabling fine scaled transmission mapping and analysis of how changes in population behaviour impact patterns of transmission.^4^ For example, genomics has been successfully used to disentangle the timeline of mass spread in Europe,^5^ and to understand community transmission versus international importations in New Zealand.^6^ Despite hundreds of SARS-CoV-2 genomes having been made available from Bangladesh, only one study so far has described the phylogenetic placement of one strain in the global context.^7^ These phylogenetic approaches provide a window into the biology of transmission that is not dependent on health system capacity to detect cases. Likewise, mobile phone data has also been used extensively as a way to monitor the population behavioural response to the epidemic in real-time, and to understand the human drivers of transmission.^8^ These new data streams may be particularly powerful tools for monitoring the pandemic in LMICs, where RT-PCR testing capacity can be often highly constrained.

Here we combined viral genomic and population mobility data to analyse the emergence and outbreak trajectory of SARS-CoV-2 in Bangladesh. We sequenced 67 selected viral genomes from six administrative areas (divisions) of Bangladesh using a nanopore MinION device and combined this sequence data with pre-deposited Bangladeshi and global genomes to show how early repeated international introductions into Bangladesh were replaced by endemic spread of three dominant introduced lineages dispersing across the country in late March. Population mobility patterns analyzed from digital trace and mobile phone data showed that the switch in the genomic dynamics of SARS-CoV-2 coincided with migration out of cities to the rest of the country.

## Materials and methods

### Sample Collection

As of December 21, 2020, the total number of COVID-19 testing facility across Bangladesh was 113,^3^ IEDCR was the first institute in the country to start testing SARS-CoV-2 by RT-PCR. Thus, samples received at IEDCR were available for sequencing from the start of the outbreak. In total, we sequenced 67 COVID-19 positive samples between March 8 and July 5, 2020. Additionally, we included 324 Bangladeshi and 68,870 global SARS-CoV-2 sequences in our analysis that had been deposited in GISAID (Global Initiative on Sharing All Influenza Data) as of July 31, 2020.

### Whole-genome sequencing

Patient samples that were positive for SARS-CoV-2 by RT-PCR were selected for sequencing. Stored nasopharyngeal swabs were re-extracted by QIAamp Viral RNA Mini Kit (QIAGEN) and confirmed by RT-PCR using WHO recommended primers and probes targeting the E and N gene. Specimens with Ct values less than 31 were taken forward. We performed whole genome sequencing following the ARTIC network protocol v2 (https://www.protocols.io/view/ncov-2019-sequencing-protocol-v2-bdp7i5rn?version_warning=no). Briefly, viral cDNA was synthesized with SuperScript IV (ThermoFisher, Waltham, MA, USA), followed by the second-strand synthesis with the Q5 high-fidelity DNA polymerase (New England BioLabs, USA). Sequence libraries were then constructed using the Oxford Nanopore ligation sequencing kit (SQK-LSK109). Libraries were sequenced using R9.4.1 MinION flow cells. Approximately 30 ng of final library DNA was loaded onto the flow cell and run for 18 hours. Sequenced genomes were recovered by mapping reads against the reference Wuhan genome (GenBank accession no. MN908947.3) with the ARTIC medaka (ARTIC-nCoV-bioinformaticsSOP-v1.1.0) pipeline normalised to 200X coverage for single nucleotide polymorphism (SNP) calling and generation of consensus sequence. Lineages were assigned according to the proposed nomenclature of pangolin lineage assignment software v2.0.7 and lineage version 2020-08-29 (https://github.com/hCoV-2019/pangolin).

### Phylogenetic analysis

SARS-CoV-2 sequences from Bangladesh (n=391) were phylogenetically placed on a global tree of 68,870 SARS-CoV-2 global sequences available from GISAID (July 31, 2020). The total available on July 31, 2020 (78,448) were filtered for those with short/ambiguous sequences to leave 68,870 sequences. The global tree was provided by Rob Lanfear in his 31/7/20 update.^9^ The alignment of 68,870 sequences and global tree were used with llama software version 0.1 to place the 324 Bangladeshi sequenced strains in global phylogenetic context as visualised in figure 1A.^10^ Llama parameters including a selection of five lineage representatives and an extraction of six from the larger tree radius were used to investigate the phylogeography of the Bangladesh isolates.

**Figure 1.**
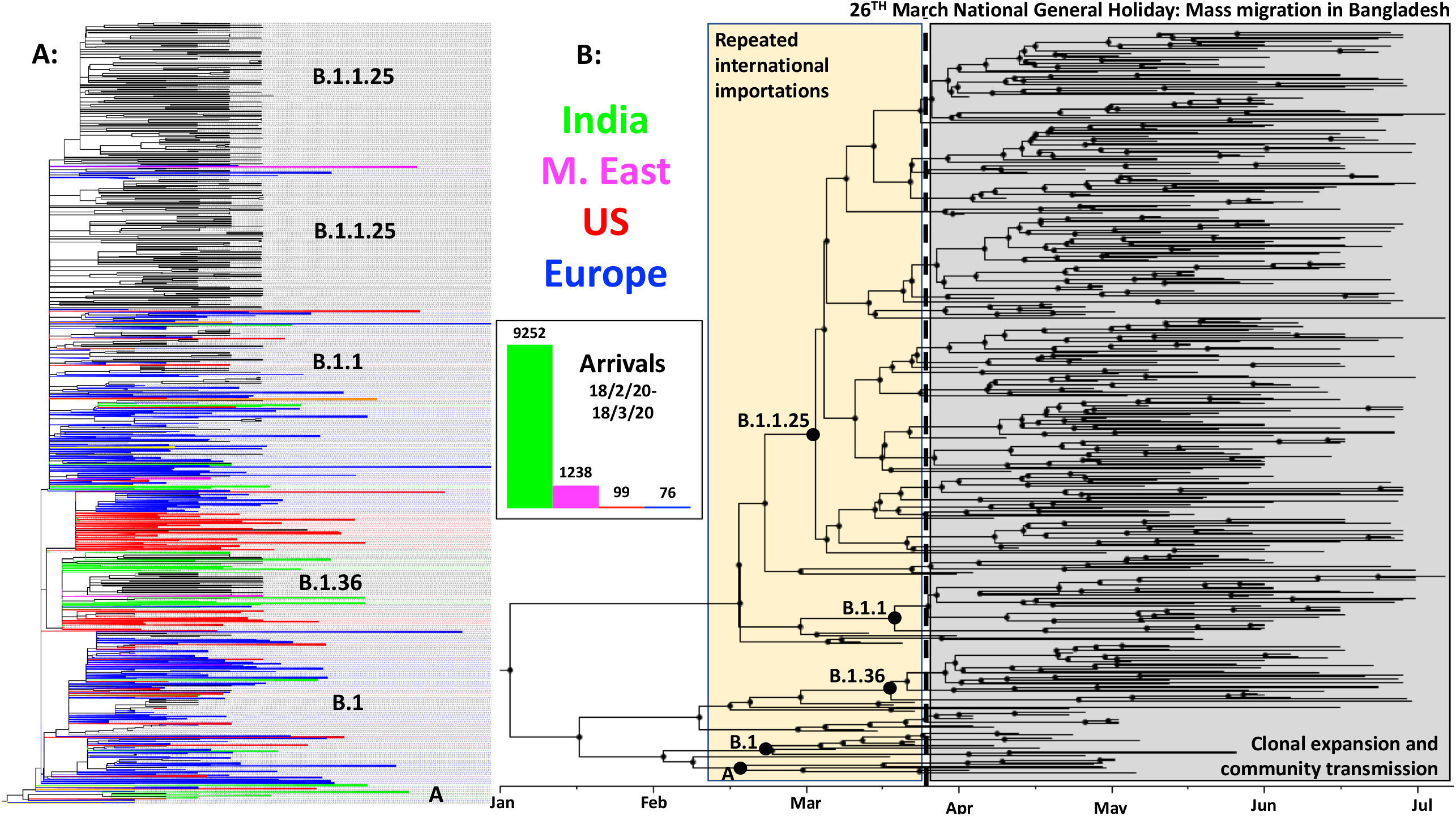
**A**. Maximum-likelihood phylogenetic analysis of 391 viruses sampled from Bangladesh (black) on a background of 68,870 publicly available GISAID global sequences in collapsed clades. Collapsed clades are coloured by the predominant region/country that they are sampled in, blue = Europe, red = US, green = India and pink = M. East. International arrival frequencies from those regions between 18/2/20-18/3/20 are displayed in a bar chart and coloured in the same colours. **B**. Maximum-likelihood Bayesian time-scaled phylogenetic analysis generated with BEAST (v 1.10.4) with clades, internal nodes and migration events annotated.

With the full set of Bangladesh sequences, we used a time-aware coalescent Bayesian exponential growth model available in BEAST (v 1.10.4). The HKY+Γ model of nucleotide substitution was used with a strict molecular clock. Parameters were estimated using Bayesian Markov Chain Monte Carlo (MCMC) framework, with 100,000,000 steps-long chains, sampling every 1000 steps and removing the initial 10% as burn-in. Sufficient sampling was assessed using Tracer (v 1.7.1), by verifying that every parameter had effective sampling sizes above 100. The resulting phylogenies were visualized with a maximum clade credibility tree in FigTree, v1.4.4.

### Mobility analysis

Anonymized and aggregated daily population location data of Facebook users in Bangladesh was provided by Facebook Data for Good.^11^ This data captures Facebook users that are providing location information through the Facebook app by having location services enabled. The population of Facebook users is determined by the modal location for each individual during every 8-hour time window. The data analyzed here was aggregated temporally (daily) and spatially (districts) and represents the daily count of Facebook users in each district from March 22 onwards. We compare these counts to the average number of users in each location during a 45-day baseline period preceding March 22. The baseline averages are calculated for each unique day of week and time of day combination. For each district, we calculated the daily percentage change in Facebook users compared to the corresponding baseline average.

We derived population mobility estimates from mobile phone call detail records (CDR) provided by three of the four telecommunication operators (Grameenphone, Banglalink, Robi Axiata Limited) in Bangladesh. Trips were calculated based on changes in a subscriber’s assigned tower location from the previous day. All data were aggregated temporally (daily) and spatially (Upazilas; sub-district) based on tower locations, following previously described methods.^8,12,13^ The aggregated data consists of the daily number of subscribers for each Upazila and the total number of trips between all pairs of Upazilas from April 27 onwards. While the Facebook data provides greater temporal coverage, it represents a much smaller percentage of the Bangladeshi population. The mobile phone data represents around 100 million subscribers across Bangladesh and allows us to estimate the daily each month, including the Eid holiday period, at the end of May 2020.

## Results

### Genomic analysis reveals multiple SARS-CoV-2 lineages circulating in Bangladesh

SARS-CoV-2 positive samples were collected by IEDCR from patients in six divisions between March and July (Supplementary Table 1). Samples were tested for SARS-CoV-2 using RT-PCR and 67 positive samples were transferred to ideSHi for sequencing. We combined our data with 324 publicly available Bangladeshi genomes (Supplementary Table 2) and also added 68,870 international contextual genomes taken from GISAID (see figure 1A).

The 391 Bangladeshi isolates fell into 19 lineages, assigned by Pangolin lineage software (lineages A, A.9, B, B.1, B.1.1, B.1.1.1, B.1.1.25, B.1.1.25.2, B.1.1.59, B.1.1.60, B.1.148, B.1.159, B.1.2, B.1.36, B.1.5, B.1.5.12, B.1.79, B.1.93 and B.2.1) (Supplementary Table 3). Of the 19 lineages, 85% of isolates fell into the three dominant lineages all possessing SNVs A23403G, C14408T, C3037T, see supplementary table 4 for a breakdown of non-synonymous SNVs in coding regions. Bayesian phylodynamic analysis estimated the mutation rate of the Bangladeshi isolates to 0·7 x 10^−3^ subs/site/year (∼20 mutations per genome per year) which is consistent with global estimates. Phylodynamic analysis of the most recent common ancestor (MRCA) predicted that SARS-CoV 2 first appeared in mid-February in Bangladesh (see figure 1B), consistent with other global estimates.^5^ The first diagnostic positive case was rapidly detected in early March, within 2 weeks of the predicted first introduction.

### Population dynamics of SARS CoV-2 across Bangladesh

From analysis of all 391 available Bangladeshi sequences, it is apparent that Pangolin Lineage B.1 was the first SARS-CoV-2 lineage observed in Bangladesh; taken on March 7 from a traveller returning from Italy. Whilst B.1 predominated until the end of March, multiple other minor lineages had also been introduced (B.1.5.12, B.2.1, B.1.2, B.1.148 and B.1.79) during this month but failed to disseminate widely. After March three lineages dominated, Lineages B.1.1 and B.1.1.25 first seen in early April, 5/4/20 and 8/4/20 respectively, and then frequently after that and lineage B.1.36, first seen at the end of March. None of these defined lineages shared a MRCA with other lineages only present in Bangladesh, rather they were all derived from lineages established and circulating outside of Bangladesh and so represent separate introductions from the global outbreak pool. B.1.1.25 has also been observed in the UK and Australia. Our data suggests that Lineage B.1.1.25 was imported into Bangladesh at least twice prior to the cessation of international air travel on March 21 (figure 1A; figure 1B). B.1.1 appeared to have been imported into Bangladesh at least five times from both the US and Europe (figure 1A). However, unlike lineages B.1.1 and B.1.1.25, phylogenetic analysis of lineage B.1.36 indicates a single importation event linked to a returning traveller from Saudi Arabia who tested positive on March 22 in Chattogram division.

All three dominant lineages expanded clonally through sustained community transmission after the end of March (figure 1B) representing 19% (B.1.1), 58% (B.1.1.25) and 8% (B.1.36) of all samples sequenced after this time. Lineage B.1.1 and B.1.1.25 were found dispersed throughout all the eight divisions of Bangladesh. Lineage B.1.36 predominated in southern Bangladesh with 64% of isolates found in Chattogram division. The remaining B.1.36 samples were found in Dhaka, Barisal, Khulna, Rajshahi, Rangpur and Sylhet but only in small numbers seen in up to two samples at each listed location.

Interestingly Lineage A, associated with the very beginning of the pandemic in China in 2019 before the introduction into Europe in 2020 was not sampled in Bangladesh until April (figure 1B), it did not expand or disseminate widely, and could have been introduced through its widespread dispersal in India before appearing in Bangladesh (figure 1A).

### Mobility data analysis reveals a mass migration in Bangladesh at the end of March

Mobility data from Facebook users was available from March 22, and from mobile phone operators from April 27. Analysis of the mobility patterns among Facebook users (figure 2A) shows that mass migration out of Dhaka occurred to all areas of the country between March 23^rd^ and March 26^th^. The displacement of individuals from the city to other parts of the country is illustrated by the increase in population relative to baseline in rural areas like Barisal. Note the large spike of movement associated with the evacuation of Barisal due to Cyclone Amphan in May during mass evacuations.^14^ Interestingly, from figure 2A its apparent that in areas like Gazipur and Narayanganj following an initial displacement, people appeared to have returned in late April;^15^ our data shows that this was followed by a more general return to Dhaka in late May (figure 2A).

**Figure 2.**
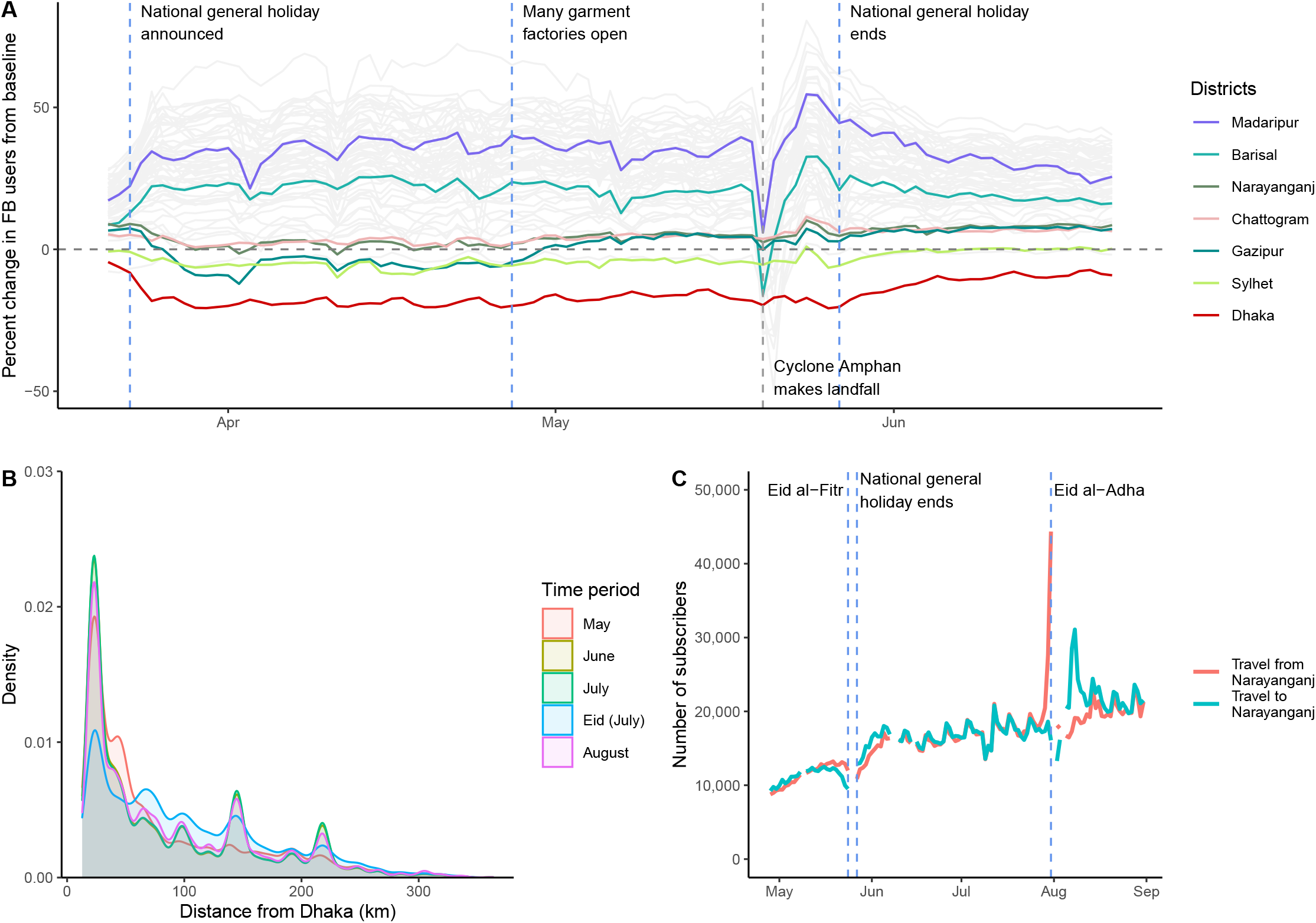
**A**. Percentage change in Facebook user population over time for each district in Bangladesh, compared to a baseline average. Specific districts are highlighted; Dhaka district is shown in red. Dashed vertical lines indicate notable events. **B**. Distribution of trips from Dhaka by distance (kilometres) travelled for each month and during Eid (July 29-31) based on CDR data from three mobile phone operators. **C**. Number of subscribers traveling to and from Narayanganj.

Figure 2B illustrates the distribution of journey lengths taken by all subscribers from three of four Bangladeshi operators from May to August, which reflects movement patterns of approximately 100 million subscribers. The impact of Eid – a national holiday during which many people travel to visit family and friends – is clearly visible in July, with more long-distance trips occurring compared to other months. In general, the distances travelled are considerable given the country’s geography (Bangladesh spans 600km east to west) and are consistent with Bangladesh’s highly mobile populations. Figure 2C shows the large number of subscribers traveling to and from Narayanganj, one of the first hotspots of SARS-CoV-2 and a continuing driver of transmission, throughout the summer. Together, these mobility data are consistent with the rapid dissemination of SARS-CoV-2 out of Dhaka to the rest of the country as people left the city at the end of March, and frequent travel around Bangladesh sustaining transmission subsequently.

### Combining time series genomic data with population mobility data

Combining the two data streams from population mobility data and genomic data, reveals a significant link between the expansion of three main lineages and the mass migration at the end of March and the beginning of the general holiday. This is evidenced by the maximum clade credibility tree in figure 1B that dates the MRCA for all three dominant lineages directly before the observed mass migration and following that the mobility data fluctuations in data usage swap from cities to regional towns (figure 2).

Given the daily migration of the Bangladeshi population into and out of major cities, the risk for sustained transmission and expansion of SARS-CoV-2 remains extremely high and limits the effectiveness of behavioural interventions. This highly mobile population appears to have rapidly transported lineages across the country, initially linked to workers living and working in cities returning home to rural areas when schools, offices and other working places were closed.

## Discussion

Our analyses of the genomic epidemiology of SARS-CoV-2 and population mobility in Bangladesh indicate that repeated international importations until late March were followed by a period of sustained community transmission consistent with a mass exodus from urban areas. A ban on international and domestic flights (March 21^st^) was likely to be the end point of international importations and March 23^rd^ was the starting point of expansion and dispersal of three dominant lineages across all of Bangladesh. The predicted introduction of SARS-CoV-2 in Bangladesh in mid-February is consistent with other global estimates that predicted lineage B.1 was imported into Italy in late January but not detected until mid-February.^4^

Recent studies have linked a ∼50 kb genomic locus inherited by modern humans from Neanderthals to heightened risk of severe disease from COVID-19.^16,17^ Importantly, the highest carrier frequency of this high-risk haplotype is found in Bangladesh underlining the need for the continued strengthening of existing surveillance, monitoring systems and interventions to reduce transmission. Novel evidence from this study indicates that non-pharmaceutical interventions that have been successful in high income countries (HIC) (e.g. stay at home orders) can exacerbate transmission in LMICs with similar demographic specificities to Bangladesh.

There are a number of limitations to our study that should be considered in the context of the results. The number of sequences sampled represented a small fraction of the documented number of positive cases at the time in Bangladesh (0·1%); the sample was selected to be a country-wide representative given limited resources for genome sequencing and consequently, the introduced viral diversity may also have been underestimated. However, the mutation rate of SARS-CoV-2 is slow (our estimate from Bangladeshi samples is 0·7 x 10^−3^ subs/site/year and latest global estimates are similar at 0·8 x 10^−3^ subs/site/year) so it would be expected that multiple studies sampling widely across all areas of Bangladesh would sample the limited diversity of lineages present in a country isolated by a ban on international arrivals.

Despite these limitations, the study is the first of its kind and exemplifies the unique advantage of combining mobility and genomic data to untangle outbreak dynamics during an outbreak. Here we provide support for a more comprehensive country-wide study of the epidemiology and spread of SARS-CoV-2, with the integration of population mobility data, in settings like Bangladesh. Recent evidence of variants of interest emerging in the UK and South Africa have heightened calls for systematic genomic surveillance in more countries globally^18^. None of the variants of interest have yet been identified in Bangladesh (as of 4/1/21) but increased vigilance and surveillance is extremely necessary.

Thus far all studies that have discussed the effect of non-pharmaceutical interventions on lineage diversity have focussed on HIC settings.^19^ Whilst few studies have effectively investigated the intricacies and specificities of differences in the dissemination of SARS-CoV-2 in low income settings, those that have, revealed that dogma on transmission dynamics can be completely different from HIC settings.^20^ Thus calling for situation appropriate tailor-made policies for all settings. It is important that the lessons learnt from Bangladesh are shared, so that other countries with similar healthcare challenges can benefit from these experiences to help suppress transmission of SARS-CoV-2. In conclusion, based on the genomic data on the SARS-CoV-2 isolates and the successful immunization programs in Bangladesh, it appears that COVID-19 vaccines becoming available will be efficacious in Bangladesh and that ongoing genomic epidemiology is essential to monitor for variants of interest under surveillance worldwide.

## Data Availability

Data sharing
All sequence data used in this study is available on GISAID as detailed in the GISAID acknowledgements section of the supplementary material.

## Contributors

LAC assisted study planning, supervised sequencing methods, performed data analysis, figure generation, and manuscript writing. MHA assisted study planning, supervised sample processing, performed sequencing, cured metadata, contributed to data interpretation, and manuscript writing. SIAR assisted the sequencing, cured metadata, generated data tables, contributed to data interpretation, and assisted manuscript writing. MMAM undertook sample processing and collected metadata. TC, AM collected and analysed mobility data and contributed to manuscript writing. MMB, MHK, SS, TK, NB, and ANA collected samples and metadata. MZR, KM, SB, reviewed the manuscript. AC collected mobility data. MSF reviewed the manuscript. SB assisted in sourcing reagents and reviewed the manuscript. NRT, COB, FQ, TS co-conceived the study, supervised the study analysis plan, and co-wrote the manuscript.

## Data sharing

All sequence data used in this study is available on GISAID as detailed in the GISAID acknowledgements section of the supplementary material.

## Acknowledgements

We would like to acknowledge Sally Forrest for her efforts in shipping reagents during the pandemic that enabled the sequencing detailed in this report. We would also like to acknowledge diagnostic testing staff at IEDCR and ideSHI who provided initial diagnostics of SARS-CoV-2 samples. icddr,b is grateful to the Governments of Bangladesh, Canada, Sweden and UK for providing core/unrestricted support.

## Supplementary material

Supplementary table 1. 67 SARS-CoV-2 samples sequenced and metadata of sample date, Ct value, District, Sex, Age, Travel history, Pangolin lineage and countries lineage also found in.

Supplementary table 2. 324 SARS-CoV-2 sequences sampled in Bangladesh sourced from GISAID and metadata detailed on sample date, division collected in, assigned Pangolin lineage and countries Pangolin lineage also found in.

Supplementary Table 3. Lineage distribution of 391 SARS-CoV-2 isolates in Bangladesh. Supplementary Table 4. List of non-synonymous SNP present in Bangladeshi SARS-CoV-2 isolates.

GISAID acknowledgements.

## References

1 Worldbank. Rural population (% of total population) – Bangladesh. https://data.worldbank.org/indicator/SP.RUR.TOTL.ZS?locations=BD. Date accessed: October 26, 2020

2 Asian Development Bank. Poverty Data: Bangladesh. https://www.adb.org/countries/bangladesh/poverty. Date accessed October 26,2020.

3 WHO report, Situation report – 43, Dec 21. https://www.who.int/docs/default-source/searo/bangladesh/covid-19-who-bangladesh-situation-reports/who_covid-19-update_43_20201221.pdf?sfvrsn=30e72858_9. Date accessed: January 4, 2021.

4 Rambault A, Holmes EC, O’Toole A et al. A dynamic nomenclature proposal for SARS-CoV-2 lineages to assist genomic epidemiology. Nat Microbiol. 2020; 5:1403–1407. doi:10.1038/s41564-020-0770-5.

5 Worobey M, Pekar J, Larsen BB et al. The emergence of SARS-CoV-2 in Europe and North America. Science. 2020. doi:10.1126/science.abc8169.

6 Geoghegan J, Ren X, Storey M et al. Genomic epidemiology reveals transmission patterns and dynamics of SARS-CoV-2 in Aotearoa New Zealand. MedxRxiv. 2020. doi:10.1101/2020.08.05.20168930.

7 Saha S, Malaker R, Sajib MSI et al. Complete Genome Sequence of a Novel Coronavirus (SARS-CoV-2) Isolate from Bangladesh. Microbiol Resour Announc. 2020. 11;9(24):e00568–20. doi:10.1128/MRA.00568-20

8 Buckee CO, Engø-Monsen K. Mobile phone data for public health: towards data-sharing solutions that protect individual privacy and national security. aRxiv. 2016. arXiv:1606.00864.

9 Lanfear R. A global phylogeny of SARS-CoV-2 from GISAID data, including sequences deposited up to 31-July-2020. Zenodo. 2020. doi:10.5281/zenodo.3958883.

10 Llama software: Local Lineage and Monophyly Assessment. https://github.com/cov-lineages/llama. Date accessed: October 27 2020

11 Maas P. Facebook Disaster Maps: Aggregate Insights for Crisis Response & Recovery. In Proceedings of the 25th ACM SIGKDD International Conference on Knowledge Discovery & Data Mining - KDD ‘19(ACM Press, Anchorage, AK, USA, 2019), 3173–3173.ISBN: 978-1-4503-6201 6.

12 Wesolowski A, Zu Erbach-Schoenberg E, Tatem AJ et al. Multinational patterns of seasonal asymmetry in human movement influence infectious disease dynamics. Nature Comms. 2017;8(1). doi:10.1038/s41467-017-02064-4.

13 Wesolowski A, Qureshi T, Boni MF et al. Impact of human mobility on the emergence of dengue epidemics in Pakistan. PNAS. 2015;112(38):11887–92. doi:10.1073/pnas.1504964112.

14 BBC. Amphan: India and Bangladesh evacuate millions ahead of super cyclone. https://www.bbc.com/news/world-asia-india-52718826. Date accessed: October 26, 2020.

15 NBC Montana. Bangladesh reopens 600 apparel factories despite virus risk. https://nbcmontana.com/news/nation-world/bangladesh-reopens-600-apparel-factories-despite-virus-risk. Date accessed: October 26,2020.

16 Ellinghaus D, Degenhardt F, Bujanda L et al. Genomewide Association Study of Severe Covid-19 with Respiratory Failure. N. Engl. J. Med. 2020. doi:10.1056/NEJMoa2020283.

17 Zeberg H, Paabo,S. The major genetic risk factor for severe COVID-19 is inherited from Neanderthals. Nature. 2020. doi:10.1038/s41586-020-2818-3.

18 SARS-CoV-2 lineages, New variant report. https://cov-lineages.org/global_report.html. Date accessed: January 4, 2021.

19 du Plessis L, McCrone JT, Zarebski AE et al. Establishment & lineage dynamics of the SARS-CoV-2 epidemic in the UK. MedRxiv. 2020. doi:10.1101/2020.10.23.20218446.

20 Laxminarayan R, Wahl B, Dudula SR et al. Epidemiology and transmission dynamics of COVID-19 in two Indian states. Science. 2020. doi:10.1126/science.abd7672.

