## Supplementary material for "Genomic and mobility data reveal mass population movement as a driver of SARS-CoV-2 dissemination and diversity in Bangladesh"

**Supplementary materials.**

**Supplementary table 1**. 67 SARS-CoV-2 samples sequenced and metadata of sample date, Ct value, District, Sex, Age, Travel history, Pangolin lineage and countries lineage also found in. FDMN = Forcibly Displaced Myanmar Nationals.

| **ID** | **Sample** | **Ct** | **Division** | **District, Area** | **Sex** | **Travel History/source of transmission** | **Lineage** | **Originate** |
| --- | --- | --- | --- | --- | --- | --- | --- | --- |
| G-85 | May-20 | 17.85 | Dhaka | Dhaka | Male | Contact of COVID-19 positive, Reinfection | B.1.1 | UK, USA, Portugal |
| G-66 | Apr-20 | 29.49 | Dhaka | Narayanganj,Araihazar | NA | NA | B.1.1 | UK, USA, Portugal |
| G-40 | Mar-20 | 20.94 | Dhaka | Dhaka,Uttara | NA | NA | B.1.2 | USA, UK, Australia |
| G-38 | Mar-20 | 16.32 | Dhaka | Dhaka,Mirpur | NA | Contact of USA returnee | B.1.79 | UK, India, Cyprus |
| G-31 | Mar-20 | 16.4 | Dhaka | Dhaka | Female | Italy returnee | B.1.79 | UK, India, Cyprus |
| G-29 | Mar-20 | 22.9 | Dhaka | Dhaka,Khilgaon | NA | NA | B.1.148 | UK |
| G-27 | Mar-20 | 19.7 | Rangpur | Gaibandha | NA | Contact of Sample ID G-23 | B.1 | UK, USA, Australia |
| G-24 | Mar-20 | 28.21 | Dhaka | Madaripur | NA | Contact of Italy returnee | B.1.5.12 | Senegal |
| G-23 | Mar-20 | 21.5 | Rangpur | Gaibandha | Female | USA Returnee | B.1.2 | USA, UK, Australia |
| G-220 | May-20 | 23.08 | Barishal | Barishal | NA | NA | B.1.1 | UK, USA, Portugal |
| G-219 | May-20 | 20.15 | Barishal | Barishal | NA | NA | B.1.1 | UK, USA, Portugal |
| G-218 | May-20 | 28.05 | Barishal | Barishal | NA | NA | B.1.1 | UK, USA, Portugal |
| G-217 | May-20 | 24.55 | Barishal | Barishal | NA | NA | B.1.1 | UK, USA, Portugal |
| G-216 | May-20 | 23.01 | Barishal | Barishal | NA | NA | B.1.1 | UK, USA, Portugal |
| G-203 | May-20 | 22.01 | Barishal | Barishal | NA | NA | B.1.93 | UK, Switzerland, Spain |
| G-201 | May-20 | 24.41 | Barishal | Barishal | NA | NA | A.9 | India |
| G-194 | Jul-20 | 21.59 | Barishal | Barishal | NA | NA | B.1.1 | UK, USA, Portugal |
| G-193 | Jul-20 | 16.42 | Chattogram | Cox's Bazar | NA | NA | B.1.36 | India, Saudi_Arabia, Bangladesh |
| G-192 | Jul-20 | 21.21 | Chattogram | Cox's Bazar | NA | NA | B.1.93 | UK, Switzerland, Spain |
| G-187 | Jun-20 | 25.89 | Chattogram | Cox's Bazar,FDMN | NA | NA | B.1 | UK, USA, Australia |
| G-185 | Jun-20 | 24.8 | Chattogram | Cox's Bazar,FDMN | NA | NA | B.1.1 | UK, USA, Portugal |
| G-184 | Jun-20 | 23.06 | Chattogram | Cox's Bazar,FDMN | NA | NA | B.1 | UK, USA, Australia |
| G-183 | Jun-20 | 18.99 | Chattogram | Cox's Bazar,FDMN | NA | NA | B.1.1 | UK, USA, Portugal |
| G-182 | Jun-20 | 19.67 | Chattogram | Cox's Bazar,FDMN | NA | NA | B.1.1 | UK, USA, Portugal |
| G-176 | Jul-20 | 21.22 | Barishal | Barishal | NA | NA | B.1.1 | UK, USA, Portugal |
| G-173 | Jul-20 | 23.17 | Chattogram | Cox's Bazar | NA | NA | B.1.1.25.2 | UK |
| G-171 | Jul-20 | 18.27 | Chattogram | Cox's Bazar | NA | NA | B.1.36 | India, Saudi_Arabia, Bangladesh |
| G-168 | Jul-20 | 17.9 | Chattogram | Cox's Bazar | NA | NA | B.1.1 | UK, USA, Portugal |
| G-156 | Jun-20 | 21.96 | Chattogram | Cox's Bazar | NA | NA | B.1.36 | India, Saudi_Arabia, Bangladesh |
| G-152 | Jun-20 | 15.99 | Chattogram | Cox's Bazar | NA | NA | B.1.36 | India, Saudi_Arabia, Bangladesh |
| G-149 | Jun-20 | 18.14 | Chattogram | Cox's Bazar | NA | NA | B.1.36 | India, Saudi_Arabia, Bangladesh |
| G-144 | Jul-20 | 29 | Dhaka | Dhaka | NA | NA | B.1.1 | UK, USA, Portugal |
| G-142 | Jun-20 | 28 | Dhaka | Dhaka | NA | NA | B.1 | UK, USA, Australia |
| G-14 | Apr-20 | 26.9 | Dhaka | Narayanganj,Siddhiganj | NA | NA | B.1.1 | UK, USA, Portugal |
| G-134 | Jun-20 | 21.8 | Dhaka | Dhaka | NA | NA | B.1.1 | UK, USA, Portugal |
| G-131 | Jul-20 | 24.5 | Dhaka | Dhaka | NA | NA | B.1.1 | UK, USA, Portugal |
| G-130 | Jul-20 | 22 | Dhaka | Dhaka | NA | NA | B.1.1 | UK, USA, Portugal |
| G-129 | Jul-20 | 19 | Dhaka | Dhaka | NA | NA | B.1.1 | UK, USA, Portugal |
| G-127 | Jul-20 | 16.84 | Dhaka | Dhaka | NA | NA | B.1.1 | UK, USA, Portugal |
| G-126 | Jul-20 | 16 | Dhaka | Dhaka | NA | NA | B.1.1 | UK, USA, Portugal |
| G-123 | Jul-20 | 28.2 | Dhaka | Dhaka | NA | NA | B.1 | UK, USA, Australia |
| G-119 | Jun-20 | 24 | Dhaka | Dhaka | NA | NA | B.1.5.12 | Senegal |
| G-115 | Jun-20 | 30 | Dhaka | Dhaka | NA | NA | B.1 | UK, USA, Australia |
| G-111 | Jun-20 | 28.3 | Dhaka | Dhaka | NA | NA | B.1 | UK, USA, Australia |
| G-110 | Jun-20 | 22.2 | Dhaka | Dhaka | NA | NA | B.1 | UK, USA, Australia |
| G-11 | Apr-20 | 25.5 | Dhaka | Dhaka | NA | Contact of Italy returnee | B.1 | UK, USA, Australia |
| G-108 | Jun-20 | 26 | Dhaka | Dhaka | NA | NA | B.1 | UK, USA, Australia |
| G-106 | Jun-20 | 23.2 | Dhaka | Dhaka | NA | NA | B.1 | UK, USA, Australia |
| G-104 | Jul-20 | 23 | Dhaka | Dhaka | NA | NA | B.1.1.1 | UK, Australia, Belgium |
| G-103 | Jun-20 | 24.12 | Dhaka | Dhaka | NA | NA | B.1.1 | UK, USA, Portugal |
| G-10 | Apr-20 | 23.5 | Dhaka | Narayanganj,Siddhiganj | NA | NA | B.1.1 | UK, USA, Portugal |
| 638 | Apr-20 | 29.82 | Dhaka | Dhaka | NA | NA | B.1 | UK, USA, Australia |
| 374 | Mar-20 | 27 | Dhaka | Dhaka,Mohakhali | Male | NA | B.2.1 | UK, Australia, USA |
| 361 | Apr-20 | 26.3 | Dhaka | Narayanganj | Female | NA | B.1.1 | UK, USA, Portugal |
| 294 | Mar-20 | 23 | Dhaka | Madaripur | Female | Contact of Italy returnee | B.1 | UK, USA, Australia |
| 29156 | Apr-20 | 23.51 | Dhaka | Dhaka | NA | NA | B.1.1 | UK, USA, Portugal |
| 2802 | Mar-20 | 29.7 | Dhaka | Narayanganj | Male | Italy returnee | B.1 | UK, USA, Australia |
| 2702 | May-20 | 19.58 | Dhaka | Dhaka | Male | NA | B.1.5.12 | Senegal |
| 256 | Mar-20 | 20.39 | Dhaka | Dhaka,Bashabo | Male | NA | B.1 | UK, USA, Australia |
| 250 | Apr-20 | 26.22 | Barishal | Barishal | Female | Contact of COVID-19 positive | B.1.5.12 | Senegal |
| 203 | Apr-20 | 22.01 | Barishal | Barishal | Male | Contact of Sample ID 201 | B.1.5 | UK, Spain, USA |
| 202 | Apr-20 | 24.23 | Barishal | Barishal | Female | Contact of COVID-19 positive | B.1.5.12 | Senegal |
| 201 | Apr-20 | 24.41 | Barishal | Barishal | Male | Travel from Barishal to Keranigonj | B.1.5.12 | Senegal |
| 1757 | Mar-20 | 28.29 | Dhaka | Dhaka | NA | NA | B.1.5.12 | Senegal |
| 1138 | Apr-20 | 27.23 | Dhaka | Narayanganj | Male | NA | B.1.5.12 | Senegal |
| 110 | Apr-20 | 31.46 | Barishal | Barishal, Barguna | Male | NA | A.9 | India |
| 109879 | Apr-20 | 31.05 | Barishal | Barishal | NA | NA | B.1.1 | UK, USA, Portugal |

**Supplementary table 2**. 324 SARS-CoV-2 sequences sampled in Bangladesh sourced from GISAID and metadata detailed on sample date, division collected in, assigned Pangolin lineage and countries Pangolin lineage also found in.

| **Sample ID** | **Sample Date** | **Division** | **Lineage** | **Originate** |
| --- | --- | --- | --- | --- |
| Bangladesh/NIB-BCSIR-02/2020 | 11-May-20 | Dhaka | B | UK, China, USA |
| Bangladesh/NIB-01/2020 | 11-May-20 | Unknown | B.1.1 | UK, USA, Portugal |
| Bangladesh/NGRI-NSTU-31/2020 | 21-Jul-20 | Chattogram | B.1.36 | India, Saudi_Arabia, Bangladesh |
| Bangladesh/NGRI-NSTU-21/2020 | 20-Jul-20 | Chattogram | B.1.36 | India, Saudi_Arabia, Bangladesh |
| Bangladesh/NGRI-NSTU-18/2020 | 21-Jul-20 | Chattogram | B.1.1.25 | Bangladesh, UK, Australia |
| Bangladesh/NGRI-NSTU-16/2020 | 21-Jul-20 | Chattogram | B.1.1.25 | Bangladesh, UK, Australia |
| Bangladesh/NGRI-NSTU-06/2020 | 23-Jul-20 | Chattogram | B.1 | UK, USA, Australia |
| Bangladesh/NGRI-NSTU-05/2020 | 23-Jul-20 | Chattogram | B.1.36 | India, Saudi_Arabia, Bangladesh |
| Bangladesh/NGRI-NSTU-03/2020 | 22-Jul-20 | Chattogram | B.1.1.25 | Bangladesh, UK, Australia |
| Bangladesh/NGRI-NSTU-02/2020 | 22-Jul-20 | Chattogram | B.1.36 | India, Saudi_Arabia, Bangladesh |
| Bangladesh/JUST-GC40.86/2020 | 11-Jun-20 | Khulna | B.1 | UK, USA, Australia |
| Bangladesh/JUST-GC40.38/2020 | 11-Jun-20 | Khulna | B.1 | UK, USA, Australia |
| Bangladesh/JUST-GC15.09/2020 | 06-May-20 | Khulna | B.1 | UK, USA, Australia |
| Bangladesh/icddrb-2083/2020 | 07-Jun-20 | Unknown | B.1.1.25 | Bangladesh, UK, Australia |
| Bangladesh/DU-CARS-5586/2020 | 13-May-20 | Dhaka | B.1.1.25 | Bangladesh, UK, Australia |
| Bangladesh/DU-50768/2020 | 06-May-20 | Dhaka | B.1 | UK, USA, Australia |
| Bangladesh/DU-50761/2020 | 06-May-20 | Dhaka | B.1.1.25 | Bangladesh, UK, Australia |
| Bangladesh/DU-50758/2020 | 06-May-20 | Dhaka | B.1.1.25 | Bangladesh, UK, Australia |
| Bangladesh/DU-50740/2020 | 06-May-20 | Dhaka | B.1.1.25 | Bangladesh, UK, Australia |
| Bangladesh/DNAS-CPH-471/2020 | 28-Apr-20 | Dhaka | B.1.1.25 | Bangladesh, UK, Australia |
| Bangladesh/DNAS-CPH-467/2020 | 28-Apr-20 | Dhaka | B.1.1.25 | Bangladesh, UK, Australia |
| Bangladesh/DNAS-CPH-466/2020 | 28-Apr-20 | Dhaka | B.1 | UK, USA, Australia |
| Bangladesh/DNAS-CPH-436/2020 | 28-Apr-20 | Dhaka | B.1.1.25 | Bangladesh, UK, Australia |
| Bangladesh/DNAS-CPH-427/2020 | 28-Apr-20 | Dhaka | B.1.1.25 | Bangladesh, UK, Australia |
| Bangladesh/CHRF-0031/2020 | 16-Jun-20 | Unknown | B.1.1.25 | Bangladesh, UK, Australia |
| Bangladesh/CHRF-0030/2020 | 11-Jun-20 | Unknown | B.1.36 | India, Saudi_Arabia, Bangladesh |
| Bangladesh/CHRF-0029/2020 | 17-Jun-20 | Unknown | B.1.1.25 | Bangladesh, UK, Australia |
| Bangladesh/CHRF-0028/2020 | 18-Jun-20 | Unknown | B.1.1.25 | Bangladesh, UK, Australia |
| Bangladesh/CHRF-0027/2020 | 16-Jun-20 | Unknown | B.1.1.25 | Bangladesh, UK, Australia |
| Bangladesh/CHRF-0025/2020 | 14-Jun-20 | Unknown | B.1.1.25 | Bangladesh, UK, Australia |
| Bangladesh/CHRF-0024/2020 | 14-Jun-20 | Unknown | B.1.36 | India, Saudi_Arabia, Bangladesh |
| Bangladesh/CHRF-0023/2020 | 12-Jun-20 | Unknown | B.1.1.25 | Bangladesh, UK, Australia |
| Bangladesh/CHRF-0022/2020 | 14-Jun-20 | Unknown | B.1.1.25 | Bangladesh, UK, Australia |
| Bangladesh/CHRF-0021/2020 | 14-May-20 | Unknown | B.1 | UK, USA, Australia |
| Bangladesh/CHRF-0020/2020 | 30-May-20 | Unknown | B.1.1.25 | Bangladesh, UK, Australia |
| Bangladesh/CHRF-0019/2020 | 14-May-20 | Unknown | B.1 | UK, USA, Australia |
| Bangladesh/CHRF-0018/2020 | 08-May-20 | Unknown | B.1.1.25 | Bangladesh, UK, Australia |
| Bangladesh/CHRF-0017/2020 | 30-May-20 | Unknown | B.1.1.25 | Bangladesh, UK, Australia |
| Bangladesh/CHRF-0016/2020 | 23-May-20 | Unknown | B.1.1.25 | Bangladesh, UK, Australia |
| Bangladesh/CHRF-0015/2020 | 02-May-20 | Unknown | B.1 | UK, USA, Australia |
| Bangladesh/CHRF-0014/2020 | 21-May-20 | Unknown | B.1.1.25 | Bangladesh, UK, Australia |
| Bangladesh/CHRF-0013/2020 | 09-May-20 | Unknown | B.1.1.25 | Bangladesh, UK, Australia |
| Bangladesh/CHRF-0012/2020 | 05-May-20 | Unknown | B.1.1.25 | Bangladesh, UK, Australia |
| Bangladesh/CHRF-0011/2020 | 30-Mar-20 | Unknown | B.1 | UK, USA, Australia |
| Bangladesh/CHRF-0010/2020 | 03-May-20 | Dhaka | B.1 | UK, USA, Australia |
| Bangladesh/CHRF-0009/2020 | 08-Apr-20 | Dhaka | B.1.1.25 | Bangladesh, UK, Australia |
| Bangladesh/CHRF-0008/2020 | 16-May-20 | Dhaka | B.1.1.25 | Bangladesh, UK, Australia |
| Bangladesh/CHRF-0007/2020 | 14-May-20 | Dhaka | B.1 | UK, USA, Australia |
| Bangladesh/CHRF-0006/2020 | 17-May-20 | Chattogram | B.1.36 | India, Saudi_Arabia, Bangladesh |
| Bangladesh/CHRF-0005/2020 | 10-May-20 | Dhaka | B.1.1.25 | Bangladesh, UK, Australia |
| Bangladesh/CHRF-0004/2020 | 17-Apr-20 | Dhaka | B.1.1.25 | Bangladesh, UK, Australia |
| Bangladesh/CHRF-0003/2020 | 17-Apr-20 | Dhaka | B.1.1.25 | Bangladesh, UK, Australia |
| Bangladesh/CHRF-0002/2020 | 20-Apr-20 | Dhaka | B.1 | UK, USA, Australia |
| Bangladesh/CHRF-0001/2020 | 18-Apr-20 | Dhaka | B.1.1.25 | Bangladesh, UK, Australia |
| Bangladesh/BCSIR-NILMRC-387/2020 | 18-Jul-20 | Chattogram | B.1.1.25 | Bangladesh, UK, Australia |
| Bangladesh/BCSIR-NILMRC-386/2020 | 21-Jul-20 | Chattogram | B.1.1.25 | Bangladesh, UK, Australia |
| Bangladesh/BCSIR-NILMRC-385/2020 | 21-Jul-20 | Chattogram | B.1.1.25 | Bangladesh, UK, Australia |
| Bangladesh/BCSIR-NILMRC-384/2020 | 21-Jul-20 | Chattogram | B.1.1.25 | Bangladesh, UK, Australia |
| Bangladesh/BCSIR-NILMRC-383/2020 | 21-Jul-20 | Chattogram | B.1.1.25 | Bangladesh, UK, Australia |
| Bangladesh/BCSIR-NILMRC-382/2020 | 21-Jul-20 | Chattogram | B.1.1.25 | Bangladesh, UK, Australia |
| Bangladesh/BCSIR-NILMRC-381/2020 | 21-Jul-20 | Chattogram | B.1.36 | India, Saudi_Arabia, Bangladesh |
| Bangladesh/BCSIR-NILMRC-374/2020 | 31-Jul-20 | Dhaka | B.1.1.60 | UK, Austria, Bangladesh |
| Bangladesh/BCSIR-NILMRC-372/2020 | 31-Jul-20 | Dhaka | B.1.1 | UK, USA, Portugal |
| Bangladesh/BCSIR-NILMRC-371/2020 | 31-Jul-20 | Dhaka | B.1.1.25 | Bangladesh, UK, Australia |
| Bangladesh/BCSIR-NILMRC-370/2020 | 21-Jul-20 | Chattogram | B.1.1.25 | Bangladesh, UK, Australia |
| Bangladesh/BCSIR-NILMRC-369/2020 | 21-Jul-20 | Chattogram | B.1.1.25 | Bangladesh, UK, Australia |
| Bangladesh/BCSIR-NILMRC-368/2020 | 21-Jul-20 | Chattogram | B.1.1.25 | Bangladesh, UK, Australia |
| Bangladesh/BCSIR-NILMRC-367/2020 | 21-Jul-20 | Chattogram | B.1.1.25 | Bangladesh, UK, Australia |
| Bangladesh/BCSIR-NILMRC-365/2020 | 21-Jul-20 | Chattogram | B.1.1 | UK, USA, Portugal |
| Bangladesh/BCSIR-NILMRC-364/2020 | 24-Jul-20 | Chattogram | B.1.1.25 | Bangladesh, UK, Australia |
| Bangladesh/BCSIR-NILMRC-363/2020 | 24-Jul-20 | Chattogram | B.1.1 | UK, USA, Portugal |
| Bangladesh/BCSIR-NILMRC-362/2020 | 24-Jul-20 | Chattogram | B.1.1 | UK, USA, Portugal |
| Bangladesh/BCSIR-NILMRC-361/2020 | 24-Jul-20 | Chattogram | B.1.1 | UK, USA, Portugal |
| Bangladesh/BCSIR-NILMRC-359/2020 | 24-Jul-20 | Chattogram | B.1.1.25 | Bangladesh, UK, Australia |
| Bangladesh/BCSIR-NILMRC-358/2020 | 24-Jul-20 | Chattogram | B.1.1.25 | Bangladesh, UK, Australia |
| Bangladesh/BCSIR-NILMRC-356/2020 | 24-Jul-20 | Chattogram | B.1.1.25 | Bangladesh, UK, Australia |
| Bangladesh/BCSIR-NILMRC-354/2020 | 15-Jul-20 | Khulna | B.1.1.25 | Bangladesh, UK, Australia |
| Bangladesh/BCSIR-NILMRC-353/2020 | 15-Jul-20 | Khulna | B.1.1.25 | Bangladesh, UK, Australia |
| Bangladesh/BCSIR-NILMRC-352/2020 | 15-Jul-20 | Khulna | B.1.1.25 | Bangladesh, UK, Australia |
| Bangladesh/BCSIR-NILMRC-351/2020 | 15-Jul-20 | Khulna | B.1.36 | India, Saudi_Arabia, Bangladesh |
| Bangladesh/BCSIR-NILMRC-350/2020 | 15-Jul-20 | Khulna | B.1.1.25 | Bangladesh, UK, Australia |
| Bangladesh/BCSIR-NILMRC-349/2020 | 15-Jul-20 | Khulna | B.1.1.25 | Bangladesh, UK, Australia |
| Bangladesh/BCSIR-NILMRC-346/2020 | 15-Jul-20 | Rajshahi | B.1.1 | UK, USA, Portugal |
| Bangladesh/BCSIR-NILMRC-345/2020 | 12-Jul-20 | Rajshahi | B.1.1.25 | Bangladesh, UK, Australia |
| Bangladesh/BCSIR-NILMRC-344/2020 | 12-Jul-20 | Rajshahi | B.1.1.25 | Bangladesh, UK, Australia |
| Bangladesh/BCSIR-NILMRC-343/2020 | 15-Jul-20 | Rajshahi | B.1.36 | India, Saudi_Arabia, Bangladesh |
| Bangladesh/BCSIR-NILMRC-342/2020 | 15-Jul-20 | Rajshahi | B.1.1.25 | Bangladesh, UK, Australia |
| Bangladesh/BCSIR-NILMRC-341/2020 | 15-Jul-20 | Rajshahi | B.1.1.25 | Bangladesh, UK, Australia |
| Bangladesh/BCSIR-NILMRC-340/2020 | 15-Jul-20 | Rajshahi | B.1.1.25 | Bangladesh, UK, Australia |
| Bangladesh/BCSIR-NILMRC-339/2020 | 15-Jul-20 | Rajshahi | B.1.1.25 | Bangladesh, UK, Australia |
| Bangladesh/BCSIR-NILMRC-338/2020 | 15-Jul-20 | Rajshahi | B.1.1.25 | Bangladesh, UK, Australia |
| Bangladesh/BCSIR-NILMRC-333/2020 | 04-Jun-20 | Mymensingh | B.1.1.25 | Bangladesh, UK, Australia |
| Bangladesh/BCSIR-NILMRC-330/2020 | 03-Jun-20 | Mymensingh | B.1.1.25 | Bangladesh, UK, Australia |
| Bangladesh/BCSIR-NILMRC-329/2020 | 03-Jun-20 | Mymensingh | B.1.1 | UK, USA, Portugal |
| Bangladesh/BCSIR-NILMRC-328/2020 | 03-Jun-20 | Mymensingh | B.1.1.25 | Bangladesh, UK, Australia |
| Bangladesh/BCSIR-NILMRC-327/2020 | 03-Jun-20 | Mymensingh | B.1.1 | UK, USA, Portugal |
| Bangladesh/BCSIR-NILMRC-326/2020 | 03-Jun-20 | Mymensingh | B.1.1 | UK, USA, Portugal |
| Bangladesh/BCSIR-NILMRC-323/2020 | 03-Jun-20 | Mymensingh | B.1.1.25 | Bangladesh, UK, Australia |
| Bangladesh/BCSIR-NILMRC-322/2020 | 31-May-20 | Mymensingh | B.1.1.25 | Bangladesh, UK, Australia |
| Bangladesh/BCSIR-NILMRC-321/2020 | 31-May-20 | Mymensingh | B.1.1.25 | Bangladesh, UK, Australia |
| Bangladesh/BCSIR-NILMRC-320/2020 | 31-May-20 | Mymensingh | B.1.1.25 | Bangladesh, UK, Australia |
| Bangladesh/BCSIR-NILMRC-319/2020 | 31-May-20 | Mymensingh | B.1.1.25 | Bangladesh, UK, Australia |
| Bangladesh/BCSIR-NILMRC-318/2020 | 31-May-20 | Mymensingh | B.1.1.25 | Bangladesh, UK, Australia |
| Bangladesh/BCSIR-NILMRC-317/2020 | 31-May-20 | Mymensingh | B.1.1.25 | Bangladesh, UK, Australia |
| Bangladesh/BCSIR-NILMRC-315/2020 | 31-May-20 | Mymensingh | B.1.1.25 | Bangladesh, UK, Australia |
| Bangladesh/BCSIR-NILMRC-312/2020 | 20-Jul-20 | Sylhet | B.1.1.25 | Bangladesh, UK, Australia |
| Bangladesh/BCSIR-NILMRC-311/2020 | 19-Jul-20 | Sylhet | B.1.1 | UK, USA, Portugal |
| Bangladesh/BCSIR-NILMRC-307/2020 | 18-Jul-20 | Sylhet | B.1.1.25 | Bangladesh, UK, Australia |
| Bangladesh/BCSIR-NILMRC-306/2020 | 18-Jul-20 | Sylhet | B.1.1 | UK, USA, Portugal |
| Bangladesh/BCSIR-NILMRC-305/2020 | 18-Jul-20 | Sylhet | B.1.1 | UK, USA, Portugal |
| Bangladesh/BCSIR-NILMRC-298/2020 | 18-Jul-20 | Sylhet | B.1.1.25 | Bangladesh, UK, Australia |
| Bangladesh/BCSIR-NILMRC-297/2020 | 18-Jul-20 | Sylhet | B.1.1.25 | Bangladesh, UK, Australia |
| Bangladesh/BCSIR-NILMRC-296/2020 | 18-Jul-20 | Sylhet | B.1.1.25 | Bangladesh, UK, Australia |
| Bangladesh/BCSIR-NILMRC-295/2020 | 18-Jul-20 | Sylhet | B.1.1.25 | Bangladesh, UK, Australia |
| Bangladesh/BCSIR-NILMRC-293/2020 | 31-May-20 | Sylhet | B.1.1 | UK, USA, Portugal |
| Bangladesh/BCSIR-NILMRC-292/2020 | 17-Jul-20 | Sylhet | B.1.1.25 | Bangladesh, UK, Australia |
| Bangladesh/BCSIR-NILMRC-291/2020 | 18-Jul-20 | Sylhet | B.1.1.25 | Bangladesh, UK, Australia |
| Bangladesh/BCSIR-NILMRC-290/2020 | 14-Jul-20 | Rangpur | B.1.1.25 | Bangladesh, UK, Australia |
| Bangladesh/BCSIR-NILMRC-288/2020 | 14-Jul-20 | Rangpur | B.1.36 | India, Saudi_Arabia, Bangladesh |
| Bangladesh/BCSIR-NILMRC-287/2020 | 14-Jul-20 | Rangpur | B.1.1 | UK, USA, Portugal |
| Bangladesh/BCSIR-NILMRC-285/2020 | 21-Jul-20 | Chattogram | B.1.1.25 | Bangladesh, UK, Australia |
| Bangladesh/BCSIR-NILMRC-284/2020 | 21-Jul-20 | Chattogram | B.1.1.25 | Bangladesh, UK, Australia |
| Bangladesh/BCSIR-NILMRC-283/2020 | 21-Jul-20 | Chattogram | B.1.1.25 | Bangladesh, UK, Australia |
| Bangladesh/BCSIR-NILMRC-282/2020 | 21-Jul-20 | Chattogram | B.1.1.25 | Bangladesh, UK, Australia |
| Bangladesh/BCSIR-NILMRC-281/2020 | 21-Jul-20 | Chattogram | B.1.1.25 | Bangladesh, UK, Australia |
| Bangladesh/BCSIR-NILMRC-280/2020 | 21-Jul-20 | Chattogram | B.1.1.25 | Bangladesh, UK, Australia |
| Bangladesh/BCSIR-NILMRC-278/2020 | 19-Jul-20 | Chattogram | B.1.1.25 | Bangladesh, UK, Australia |
| Bangladesh/BCSIR-NILMRC-276/2020 | 19-Jul-20 | Chattogram | B.1.1.25 | Bangladesh, UK, Australia |
| Bangladesh/BCSIR-NILMRC-275/2020 | 19-Jul-20 | Chattogram | B.1.1.25 | Bangladesh, UK, Australia |
| Bangladesh/BCSIR-NILMRC-274/2020 | 19-Jul-20 | Chattogram | B.1.1.25 | Bangladesh, UK, Australia |
| Bangladesh/BCSIR-NILMRC-273/2020 | 19-Jul-20 | Chattogram | B.1.1 | UK, USA, Portugal |
| Bangladesh/BCSIR-NILMRC-269/2020 | 19-Jul-20 | Chattogram | B.1.1.25 | Bangladesh, UK, Australia |
| Bangladesh/BCSIR-NILMRC-268/2020 | 19-Jul-20 | Chattogram | B.1.1.25 | Bangladesh, UK, Australia |
| Bangladesh/BCSIR-NILMRC-265/2020 | 19-Jul-20 | Chattogram | B.1.1.25 | Bangladesh, UK, Australia |
| Bangladesh/BCSIR-NILMRC-264/2020 | 16-Jul-20 | Chattogram | B.1.1.25 | Bangladesh, UK, Australia |
| Bangladesh/BCSIR-NILMRC-263/2020 | 07-Jul-20 | Khulna | B.1.1 | UK, USA, Portugal |
| Bangladesh/BCSIR-NILMRC-262/2020 | 07-Jul-20 | Khulna | B.1.1 | UK, USA, Portugal |
| Bangladesh/BCSIR-NILMRC-261/2020 | 07-Jul-20 | Khulna | B.1.1.25 | Bangladesh, UK, Australia |
| Bangladesh/BCSIR-NILMRC-260/2020 | 07-Jul-20 | Khulna | B.1.1.25 | Bangladesh, UK, Australia |
| Bangladesh/BCSIR-NILMRC-259/2020 | 07-Jul-20 | Khulna | B.1.1.25 | Bangladesh, UK, Australia |
| Bangladesh/BCSIR-NILMRC-258/2020 | 07-Jul-20 | Khulna | B.1.1.25 | Bangladesh, UK, Australia |
| Bangladesh/BCSIR-NILMRC-257/2020 | 07-Jul-20 | Khulna | B.1.36 | India, Saudi_Arabia, Bangladesh |
| Bangladesh/BCSIR-NILMRC-256/2020 | 07-Jul-20 | Khulna | B.1.1.25 | Bangladesh, UK, Australia |
| Bangladesh/BCSIR-NILMRC-255/2020 | 07-Jul-20 | Khulna | B.1.1.25 | Bangladesh, UK, Australia |
| Bangladesh/BCSIR-NILMRC-254/2020 | 07-Jul-20 | Khulna | B.1.1.25 | Bangladesh, UK, Australia |
| Bangladesh/BCSIR-NILMRC-253/2020 | 07-Jul-20 | Khulna | B.1.1.25 | Bangladesh, UK, Australia |
| Bangladesh/BCSIR-NILMRC-251/2020 | 07-Jul-20 | Khulna | B.1.1.25 | Bangladesh, UK, Australia |
| Bangladesh/BCSIR-NILMRC-250/2020 | 07-Jul-20 | Khulna | B.1.1.25 | Bangladesh, UK, Australia |
| Bangladesh/BCSIR-NILMRC-249/2020 | 07-Jul-20 | Khulna | B.1.1.25 | Bangladesh, UK, Australia |
| Bangladesh/BCSIR-NILMRC-248/2020 | 07-Jul-20 | Khulna | B.1.1.25 | Bangladesh, UK, Australia |
| Bangladesh/BCSIR-NILMRC-247/2020 | 07-Jul-20 | Khulna | B.1.1.25 | Bangladesh, UK, Australia |
| Bangladesh/BCSIR-NILMRC-246/2020 | 07-Jul-20 | Khulna | B.1.1.25 | Bangladesh, UK, Australia |
| Bangladesh/BCSIR-NILMRC-245/2020 | 06-Jul-20 | Barishal | B.1.1.25 | Bangladesh, UK, Australia |
| Bangladesh/BCSIR-NILMRC-244/2020 | 06-Jul-20 | Barishal | B.1.1.25 | Bangladesh, UK, Australia |
| Bangladesh/BCSIR-NILMRC-243/2020 | 06-Jul-20 | Barishal | B.1.1.25 | Bangladesh, UK, Australia |
| Bangladesh/BCSIR-NILMRC-242/2020 | 06-Jul-20 | Barishal | B.1.1.25 | Bangladesh, UK, Australia |
| Bangladesh/BCSIR-NILMRC-241/2020 | 06-Jul-20 | Barishal | B.1.1.25 | Bangladesh, UK, Australia |
| Bangladesh/BCSIR-NILMRC-240/2020 | 06-Jul-20 | Barishal | B.1.1.25 | Bangladesh, UK, Australia |
| Bangladesh/BCSIR-NILMRC-239/2020 | 06-Jul-20 | Barishal | B.1.1.25 | Bangladesh, UK, Australia |
| Bangladesh/BCSIR-NILMRC-238/2020 | 06-Jul-20 | Barishal | B.1.1.25 | Bangladesh, UK, Australia |
| Bangladesh/BCSIR-NILMRC-237/2020 | 06-Jul-20 | Barishal | B.1.1.25 | Bangladesh, UK, Australia |
| Bangladesh/BCSIR-NILMRC-236/2020 | 06-Jul-20 | Barishal | B.1.1 | UK, USA, Portugal |
| Bangladesh/BCSIR-NILMRC-235/2020 | 06-Jul-20 | Barishal | B.1.1.25 | Bangladesh, UK, Australia |
| Bangladesh/BCSIR-NILMRC-234/2020 | 06-Jul-20 | Barishal | B.1.1 | UK, USA, Portugal |
| Bangladesh/BCSIR-NILMRC-233/2020 | 06-Jul-20 | Barishal | B.1.1 | UK, USA, Portugal |
| Bangladesh/BCSIR-NILMRC-232/2020 | 06-Jul-20 | Barishal | B.1.36 | India, Saudi_Arabia, Bangladesh |
| Bangladesh/BCSIR-NILMRC-231/2020 | 06-Jul-20 | Barishal | B.1.1.25 | Bangladesh, UK, Australia |
| Bangladesh/BCSIR-NILMRC-230/2020 | 06-Jul-20 | Barishal | B.1.1.25 | Bangladesh, UK, Australia |
| Bangladesh/BCSIR-NILMRC-229/2020 | 06-Jul-20 | Barishal | B.1.1.25 | Bangladesh, UK, Australia |
| Bangladesh/BCSIR-NILMRC-228/2020 | 06-Jul-20 | Barishal | B.1.159 | France, Bangladesh |
| Bangladesh/BCSIR-NILMRC-227/2020 | 06-Jul-20 | Barishal | B.1.1.25 | Bangladesh, UK, Australia |
| Bangladesh/BCSIR-NILMRC-226/2020 | 06-Jul-20 | Barishal | B.1.1.25 | Bangladesh, UK, Australia |
| Bangladesh/BCSIR-NILMRC-224/2020 | 06-Jul-20 | Barishal | B.1.1 | UK, USA, Portugal |
| Bangladesh/BCSIR-NILMRC-223/2020 | 06-Jul-20 | Barishal | B.1.1 | UK, USA, Portugal |
| Bangladesh/BCSIR-NILMRC-222/2020 | 06-Jul-20 | Barishal | B.1.1 | UK, USA, Portugal |
| Bangladesh/BCSIR-NILMRC-220/2020 | 24-Jun-20 | Sylhet | B.1.36 | India, Saudi_Arabia, Bangladesh |
| Bangladesh/BCSIR-NILMRC-219/2020 | 24-Jun-20 | Sylhet | B.1.1.25 | Bangladesh, UK, Australia |
| Bangladesh/BCSIR-NILMRC-218/2020 | 24-Jun-20 | Sylhet | B.1.1.25 | Bangladesh, UK, Australia |
| Bangladesh/BCSIR-NILMRC-217/2020 | 24-Jun-20 | Sylhet | B.1.1.25 | Bangladesh, UK, Australia |
| Bangladesh/BCSIR-NILMRC-216/2020 | 24-Jun-20 | Sylhet | B.1.36 | India, Saudi_Arabia, Bangladesh |
| Bangladesh/BCSIR-NILMRC-213/2020 | 14-Jun-20 | Sylhet | B.1.1 | UK, USA, Portugal |
| Bangladesh/BCSIR-NILMRC-212/2020 | 14-Jun-20 | Sylhet | B.1.1.25 | Bangladesh, UK, Australia |
| Bangladesh/BCSIR-NILMRC-211/2020 | 14-Jun-20 | Sylhet | B.1.1.25 | Bangladesh, UK, Australia |
| Bangladesh/BCSIR-NILMRC-210/2020 | 14-Jun-20 | Sylhet | B.1.1 | UK, USA, Portugal |
| Bangladesh/BCSIR-NILMRC-208/2020 | 27-Jun-20 | Chattogram | B.1.1.25 | Bangladesh, UK, Australia |
| Bangladesh/BCSIR-NILMRC-207/2020 | 27-Jun-20 | Chattogram | B.1.1.25 | Bangladesh, UK, Australia |
| Bangladesh/BCSIR-NILMRC-206/2020 | 27-Jun-20 | Chattogram | B.1.1.25 | Bangladesh, UK, Australia |
| Bangladesh/BCSIR-NILMRC-205/2020 | 26-Jun-20 | Chattogram | B.1.1.25 | Bangladesh, UK, Australia |
| Bangladesh/BCSIR-NILMRC-204/2020 | 26-Jun-20 | Chattogram | B.1.36 | India, Saudi_Arabia, Bangladesh |
| Bangladesh/BCSIR-NILMRC-203/2020 | 20-Jun-20 | Chattogram | B.1.1.25 | Bangladesh, UK, Australia |
| Bangladesh/BCSIR-NILMRC-201/2020 | 20-Jun-20 | Chattogram | B.1.1.25 | Bangladesh, UK, Australia |
| Bangladesh/BCSIR-NILMRC-199/2020 | 20-Jun-20 | Chattogram | B.1.1.25 | Bangladesh, UK, Australia |
| Bangladesh/BCSIR-NILMRC-198/2020 | 20-Jun-20 | Chattogram | B.1.1 | UK, USA, Portugal |
| Bangladesh/BCSIR-NILMRC-196/2020 | 20-Jun-20 | Chattogram | B.1.1.25 | Bangladesh, UK, Australia |
| Bangladesh/BCSIR-NILMRC-195/2020 | 20-Jun-20 | Chattogram | B.1.1.25 | Bangladesh, UK, Australia |
| Bangladesh/BCSIR-NILMRC-194/2020 | 20-Jun-20 | Chattogram | B.1.1 | UK, USA, Portugal |
| Bangladesh/BCSIR-NILMRC-193/2020 | 17-Jun-20 | Chattogram | B.1.1.25 | Bangladesh, UK, Australia |
| Bangladesh/BCSIR-NILMRC-192/2020 | 17-Jun-20 | Chattogram | B.1.1.25 | Bangladesh, UK, Australia |
| Bangladesh/BCSIR-NILMRC-190/2020 | 17-Jun-20 | Chattogram | B.1.1.59 | UK, Bangladesh |
| Bangladesh/BCSIR-NILMRC-188/2020 | 16-Jun-20 | Chattogram | B.1.1.25 | Bangladesh, UK, Australia |
| Bangladesh/BCSIR-NILMRC-187/2020 | 16-Jun-20 | Chattogram | B.1.1.25 | Bangladesh, UK, Australia |
| Bangladesh/BCSIR-NILMRC-186/2020 | 16-Jun-20 | Chattogram | B.1.36 | India, Saudi_Arabia, Bangladesh |
| Bangladesh/BCSIR-NILMRC-184/2020 | 20-Jun-20 | Rajshahi | B.1.1.25 | Bangladesh, UK, Australia |
| Bangladesh/BCSIR-NILMRC-182/2020 | 20-Jun-20 | Rajshahi | B.1.1 | UK, USA, Portugal |
| Bangladesh/BCSIR-NILMRC-181/2020 | 20-Jun-20 | Rajshahi | B.1.1.25 | Bangladesh, UK, Australia |
| Bangladesh/BCSIR-NILMRC-180/2020 | 20-Jun-20 | Rajshahi | B.1.1.25 | Bangladesh, UK, Australia |
| Bangladesh/BCSIR-NILMRC-179/2020 | 15-Jun-20 | Rajshahi | B.1.1.25 | Bangladesh, UK, Australia |
| Bangladesh/BCSIR-NILMRC-178/2020 | 15-Jun-20 | Rajshahi | B.1.1.25 | Bangladesh, UK, Australia |
| Bangladesh/BCSIR-NILMRC-177/2020 | 12-Jun-20 | Rajshahi | B.1.1.25 | Bangladesh, UK, Australia |
| Bangladesh/BCSIR-NILMRC-176/2020 | 12-Jun-20 | Rajshahi | B.1.1.25 | Bangladesh, UK, Australia |
| Bangladesh/BCSIR-NILMRC-175/2020 | 12-Jun-20 | Rajshahi | B.1.1.25 | Bangladesh, UK, Australia |
| Bangladesh/BCSIR-NILMRC-174/2020 | 12-Jun-20 | Rajshahi | B.1.1.25 | Bangladesh, UK, Australia |
| Bangladesh/BCSIR-NILMRC-173/2020 | 18-Jun-20 | Rajshahi | B.1.1.25 | Bangladesh, UK, Australia |
| Bangladesh/BCSIR-NILMRC-172/2020 | 18-Jun-20 | Rajshahi | B.1.1.25 | Bangladesh, UK, Australia |
| Bangladesh/BCSIR-NILMRC-168/2020 | 18-Jun-20 | Rajshahi | B.1.1.25 | Bangladesh, UK, Australia |
| Bangladesh/BCSIR-NILMRC-167/2020 | 18-Jun-20 | Rajshahi | B.1.1.25 | Bangladesh, UK, Australia |
| Bangladesh/BCSIR-NILMRC-165/2020 | 18-Jun-20 | Rajshahi | B.1.1 | UK, USA, Portugal |
| Bangladesh/BCSIR-NILMRC-161/2020 | 18-Jun-20 | Rajshahi | B.1.1.25 | Bangladesh, UK, Australia |
| Bangladesh/BCSIR-NILMRC-160/2020 | 18-Jun-20 | Rajshahi | B.1.1.25 | Bangladesh, UK, Australia |
| Bangladesh/BCSIR-NILMRC-158/2020 | 18-Jun-20 | Rajshahi | B.1.36 | India, Saudi_Arabia, Bangladesh |
| Bangladesh/BCSIR-NILMRC-156/2020 | 18-Jun-20 | Rajshahi | B.1.1 | UK, USA, Portugal |
| Bangladesh/BCSIR-NILMRC-155/2020 | 18-Jun-20 | Rajshahi | B.1.1.25 | Bangladesh, UK, Australia |
| Bangladesh/BCSIR-NILMRC-154/2020 | 18-Jun-20 | Rajshahi | B.1.1 | UK, USA, Portugal |
| Bangladesh/BCSIR-NILMRC-149/2020 | 18-Jun-20 | Dhaka | B.1.1.25 | Bangladesh, UK, Australia |
| Bangladesh/BCSIR-NILMRC-144/2020 | 18-Jun-20 | Dhaka | B.1.1.25 | Bangladesh, UK, Australia |
| Bangladesh/BCSIR-NILMRC-141/2020 | 18-Jun-20 | Dhaka | B.1.1.25 | Bangladesh, UK, Australia |
| Bangladesh/BCSIR-NILMRC-138/2020 | 18-Jun-20 | Dhaka | B.1.1.25 | Bangladesh, UK, Australia |
| Bangladesh/BCSIR-NILMRC-137/2020 | 18-Jun-20 | Dhaka | B.1.1.25 | Bangladesh, UK, Australia |
| Bangladesh/BCSIR-NILMRC-135/2020 | 18-Jun-20 | Dhaka | B.1.1.25 | Bangladesh, UK, Australia |
| Bangladesh/BCSIR-NILMRC-133/2020 | 18-Jun-20 | Dhaka | B.1.1.25 | Bangladesh, UK, Australia |
| Bangladesh/BCSIR-NILMRC-132/2020 | 18-Jun-20 | Dhaka | B.1.1.25 | Bangladesh, UK, Australia |
| Bangladesh/BCSIR-NILMRC-130/2020 | 17-Jun-20 | Dhaka | B.1.1 | UK, USA, Portugal |
| Bangladesh/BCSIR-NILMRC-126/2020 | 17-Jun-20 | Rajshahi | B.1.1.25 | Bangladesh, UK, Australia |
| Bangladesh/BCSIR-NILMRC-118a/2020 | 18-Jun-20 | Dhaka | B.1.1 | UK, USA, Portugal |
| Bangladesh/BCSIR-NILMRC-118/2020 | 18-Jun-20 | Dhaka | B.1.1 | UK, USA, Portugal |
| Bangladesh/BCSIR-NILMRC-117/2020 | 07-Jun-20 | Rangpur | B.1.1 | UK, USA, Portugal |
| Bangladesh/BCSIR-NILMRC-116/2020 | 07-Jun-20 | Rangpur | B.1.1.25 | Bangladesh, UK, Australia |
| Bangladesh/BCSIR-NILMRC-115/2020 | 07-Jun-20 | Rangpur | B.1.1.25 | Bangladesh, UK, Australia |
| Bangladesh/BCSIR-NILMRC-114/2020 | 07-Jun-20 | Rangpur | B.1.1 | UK, USA, Portugal |
| Bangladesh/BCSIR-NILMRC-113/2020 | 07-Jun-20 | Rangpur | B.1.1.25 | Bangladesh, UK, Australia |
| Bangladesh/BCSIR-NILMRC-112/2020 | 07-Jun-20 | Rangpur | B.1.1 | UK, USA, Portugal |
| Bangladesh/BCSIR-NILMRC-111/2020 | 07-Jun-20 | Rangpur | B.1.1.25 | Bangladesh, UK, Australia |
| Bangladesh/BCSIR-NILMRC-110/2020 | 07-Jun-20 | Rangpur | B.1.1.25 | Bangladesh, UK, Australia |
| Bangladesh/BCSIR-NILMRC-109/2020 | 07-Jun-20 | Rangpur | B.1.1 | UK, USA, Portugal |
| Bangladesh/BCSIR-NILMRC-108/2020 | 07-Jun-20 | Rangpur | B.1.1.25 | Bangladesh, UK, Australia |
| Bangladesh/BCSIR-NILMRC-107/2020 | 07-Jun-20 | Rangpur | B.1.1.25 | Bangladesh, UK, Australia |
| Bangladesh/BCSIR-NILMRC-105/2020 | 07-Jun-20 | Rangpur | B.1.1.25 | Bangladesh, UK, Australia |
| Bangladesh/BCSIR-NILMRC-104/2020 | 07-Jun-20 | Rangpur | B.1.1.25 | Bangladesh, UK, Australia |
| Bangladesh/BCSIR-NILMRC-103/2020 | 07-Jun-20 | Rangpur | B.1.1.25 | Bangladesh, UK, Australia |
| Bangladesh/BCSIR-NILMRC-102/2020 | 07-Jun-20 | Rangpur | B.1.1.25 | Bangladesh, UK, Australia |
| Bangladesh/BCSIR-NILMRC-101/2020 | 07-Jun-20 | Rangpur | B.1.1.25 | Bangladesh, UK, Australia |
| Bangladesh/BCSIR-NILMRC-100/2020 | 23-May-20 | Rangpur | B.1.1.25 | Bangladesh, UK, Australia |
| Bangladesh/BCSIR-NILMRC-096/2020 | 23-May-20 | Rangpur | B.1.1.25 | Bangladesh, UK, Australia |
| Bangladesh/BCSIR-NILMRC-095/2020 | 23-May-20 | Rangpur | B.1.1.25 | Bangladesh, UK, Australia |
| Bangladesh/BCSIR-NILMRC-094/2020 | 23-May-20 | Rangpur | B.1.1.25 | Bangladesh, UK, Australia |
| Bangladesh/BCSIR-NILMRC-091/2020 | 02-Jun-20 | Rangpur | B.1.1.25 | Bangladesh, UK, Australia |
| Bangladesh/BCSIR-NILMRC-090/2020 | 02-Jun-20 | Rangpur | B.1.1.25 | Bangladesh, UK, Australia |
| Bangladesh/BCSIR-NILMRC-088/2020 | 09-May-20 | Dhaka | B.1.1.25 | Bangladesh, UK, Australia |
| Bangladesh/BCSIR-NILMRC-087/2020 | 09-May-20 | Dhaka | B.1.1.25 | Bangladesh, UK, Australia |
| Bangladesh/BCSIR-NILMRC-086/2020 | 01-Jun-20 | Dhaka | B.1.1 | UK, USA, Portugal |
| Bangladesh/BCSIR-NILMRC-085/2020 | 01-Jun-20 | Dhaka | B.1.1.25 | Bangladesh, UK, Australia |
| Bangladesh/BCSIR-NILMRC-084/2020 | 01-Jun-20 | Dhaka | B.1.1 | UK, USA, Portugal |
| Bangladesh/BCSIR-NILMRC-083/2020 | 01-Jun-20 | Dhaka | B.1.1.25 | Bangladesh, UK, Australia |
| Bangladesh/BCSIR-NILMRC-082/2020 | 01-Jun-20 | Dhaka | B.1.1.25 | Bangladesh, UK, Australia |
| Bangladesh/BCSIR-NILMRC-081/2020 | 01-Jun-20 | Dhaka | B.1.1.25 | Bangladesh, UK, Australia |
| Bangladesh/BCSIR-NILMRC-080/2020 | 01-Jun-20 | Dhaka | B.1.1 | UK, USA, Portugal |
| Bangladesh/BCSIR-NILMRC-078/2020 | 01-Jun-20 | Dhaka | B.1.1 | UK, USA, Portugal |
| Bangladesh/BCSIR-NILMRC-077/2020 | 01-Jun-20 | Dhaka | B.1.1.25 | Bangladesh, UK, Australia |
| Bangladesh/BCSIR-NILMRC-076/2020 | 01-Jun-20 | Dhaka | B.1.1.25 | Bangladesh, UK, Australia |
| Bangladesh/BCSIR-NILMRC-075/2020 | 01-Jun-20 | Dhaka | B.1.1.25 | Bangladesh, UK, Australia |
| Bangladesh/BCSIR-NILMRC-074/2020 | 01-Jun-20 | Dhaka | B.1.1.25 | Bangladesh, UK, Australia |
| Bangladesh/BCSIR-NILMRC-073/2020 | 01-Jun-20 | Dhaka | B.1.1.25 | Bangladesh, UK, Australia |
| Bangladesh/BCSIR-NILMRC-072/2020 | 26-May-20 | Chattogram | B.1.1.25 | Bangladesh, UK, Australia |
| Bangladesh/BCSIR-NILMRC-071/2020 | 26-May-20 | Chattogram | B.1.36 | India, Saudi_Arabia, Bangladesh |
| Bangladesh/BCSIR-NILMRC-070/2020 | 26-May-20 | Chattogram | B.1.36 | India, Saudi_Arabia, Bangladesh |
| Bangladesh/BCSIR-NILMRC-069/2020 | 26-May-20 | Chattogram | B.1.36 | India, Saudi_Arabia, Bangladesh |
| Bangladesh/BCSIR-NILMRC-067/2020 | 26-May-20 | Chattogram | B.1.36 | India, Saudi_Arabia, Bangladesh |
| Bangladesh/BCSIR-NILMRC-066/2020 | 26-May-20 | Chattogram | B.1.1.25 | Bangladesh, UK, Australia |
| Bangladesh/BCSIR-NILMRC-064/2020 | 31-May-20 | Dhaka | B.1.1.25 | Bangladesh, UK, Australia |
| Bangladesh/BCSIR-NILMRC-062/2020 | 31-May-20 | Dhaka | B.1.1.25 | Bangladesh, UK, Australia |
| Bangladesh/BCSIR-NILMRC-060/2020 | 10-May-20 | Dhaka | B.1.1.25 | Bangladesh, UK, Australia |
| Bangladesh/BCSIR-NILMRC-059/2020 | 10-May-20 | Dhaka | B.1.1.25 | Bangladesh, UK, Australia |
| Bangladesh/BCSIR-NILMRC-058/2020 | 10-May-20 | Dhaka | B.1.1.25 | Bangladesh, UK, Australia |
| Bangladesh/BCSIR-NILMRC-057/2020 | 10-May-20 | Dhaka | B.1.1 | UK, USA, Portugal |
| Bangladesh/BCSIR-NILMRC-056/2020 | 10-May-20 | Dhaka | B.1.1.25 | Bangladesh, UK, Australia |
| Bangladesh/BCSIR-NILMRC-054/2020 | 01-Jun-20 | Dhaka | B.1.1.25 | Bangladesh, UK, Australia |
| Bangladesh/BCSIR-NILMRC-053/2020 | 07-May-20 | Dhaka | B.1.1.25 | Bangladesh, UK, Australia |
| Bangladesh/BCSIR-NILMRC-052/2020 | 01-Jun-20 | Dhaka | B.1.1 | UK, USA, Portugal |
| Bangladesh/BCSIR-NILMRC-051/2020 | 07-May-20 | Dhaka | B.1.1.25 | Bangladesh, UK, Australia |
| Bangladesh/BCSIR-NILMRC-050/2020 | 21-May-20 | Dhaka | B.1.1.25 | Bangladesh, UK, Australia |
| Bangladesh/BCSIR-NILMRC-042/2020 | 23-May-20 | Dhaka | B.1.1.25 | Bangladesh, UK, Australia |
| Bangladesh/BCSIR-NILMRC-025/2020 | 24-May-20 | Dhaka | B.1.1 | UK, USA, Portugal |
| Bangladesh/BCSIR-NILMRC-021/2020 | 24-May-20 | Dhaka | B.1.1.25 | Bangladesh, UK, Australia |
| Bangladesh/BCSIR-NILMRC-018/2020 | 24-May-20 | Dhaka | B.1.1 | UK, USA, Portugal |
| Bangladesh/BCSIR-NILMRC-017/2020 | 24-May-20 | Dhaka | B.1.1.25 | Bangladesh, UK, Australia |
| Bangladesh/BCSIR-NILMRC-015/2020 | 23-May-20 | Dhaka | B.1.1.25 | Bangladesh, UK, Australia |
| Bangladesh/BCSIR-NILMRC-009/2020 | 23-May-20 | Dhaka | B.1.1.25 | Bangladesh, UK, Australia |
| Bangladesh/BCSIR-NILMRC-008/2020 | 21-May-20 | Dhaka | B.1.1.25 | Bangladesh, UK, Australia |
| Bangladesh/BCSIR-NILMRC-007/2020 | 21-May-20 | Dhaka | B.1.1 | UK, USA, Portugal |
| Bangladesh/BCSIR-NILMRC-006/2020 | 21-May-20 | Dhaka | B.1.1.25 | Bangladesh, UK, Australia |
| Bangladesh/BCSIR-NILMRC-006.2/2020 | 23-May-20 | Dhaka | B.1.1.25 | Bangladesh, UK, Australia |
| Bangladesh/BCSIR-NILMRC-005/2020 | 23-May-20 | Dhaka | B.1.1.25 | Bangladesh, UK, Australia |
| Bangladesh/BCSIR-NILMRC-004-2/2020 | 21-May-20 | Dhaka | B.1.1.25 | Bangladesh, UK, Australia |
| Bangladesh/BCSIR-NILMRC-004/2020 | 23-May-20 | Dhaka | B.1.1.25 | Bangladesh, UK, Australia |
| Bangladesh/BCSIR-NILMRC-003/2020 | 23-May-20 | Dhaka | B.1.1.25 | Bangladesh, UK, Australia |
| Bangladesh/BCSIR-NILMRC-002/2020 | 23-May-20 | Dhaka | B.1.1.25 | Bangladesh, UK, Australia |
| Bangladesh/BCSIR-NILMRC_153/2020 | 18-Jun-20 | Rajshahi | B.1.1.25 | Bangladesh, UK, Australia |
| Bangladesh/BCSIR-NILMRC_152/2020 | 18-Jun-20 | Dhaka | B.1.1.25 | Bangladesh, UK, Australia |
| Bangladesh/BCSIR-NILMRC_151/2020 | 18-Jun-20 | Dhaka | B.1.1 | UK, USA, Portugal |
| Bangladesh/BCSIR-NILMRC_150/2020 | 18-Jun-20 | Dhaka | B.1.1.25 | Bangladesh, UK, Australia |
| Bangladesh/BCSIR-NILMRC_146/2020 | 18-Jun-20 | Dhaka | B.1.1.25 | Bangladesh, UK, Australia |
| Bangladesh/BCSIR-NILMRC_145/2020 | 18-Jun-20 | Dhaka | B.1.1.25 | Bangladesh, UK, Australia |
| Bangladesh/BCSIR-NILMRC_139/2020 | 18-Jun-20 | Dhaka | B.1.1.25 | Bangladesh, UK, Australia |
| Bangladesh/BCSIR-NILMRC_131/2020 | 18-Jun-20 | Dhaka | B.1.1 | UK, USA, Portugal |
| Bangladesh/BCSIR-NILMRC_125/2020 | 17-Jun-20 | Rajshahi | B.1.1.25 | Bangladesh, UK, Australia |
| Bangladesh/BCSIR-DU-16/2020 | 12-Jul-20 | Dhaka | B.1.36 | India, Saudi_Arabia, Bangladesh |
| Bangladesh/BARJ-CVASU-CTG-518/2020 | 10-May-20 | Chattogram | A | China, India, Japan |
| Bangladesh/BARJ-CVASU-CTG-517/2020 | 03-May-20 | Chattogram | B.1.36 | India, Saudi_Arabia, Bangladesh |
| Bangladesh/BARJ-CVASU-CTG-511/2020 | 09-May-20 | Chattogram | B.1.36 | India, Saudi_Arabia, Bangladesh |
| Bangladesh/BARJ-CVASU-CTG-506/2020 | 08-May-20 | Chattogram | A | China, India, Japan |
| Bangladesh/BARJ-CVASU-CTG-503/2020 | 13-May-20 | Chattogram | A | China, India, Japan |
| Bangladesh/BARJ-CVASU-CTG-502/2020 | 13-May-20 | Chattogram | A | China, India, Japan |
| Bangladesh/BARJ-CVASU-CTG-501/2020 | 10-May-20 | Chattogram | A | China, India, Japan |
| Bangladesh/Akbiomed-01/2020 | 25-Apr-20 | Unknown | B.1 | UK, USA, Australia |

**Supplementary Table 3.** Lineage distribution of 391 SARS-CoV-2 isolates in Bangladesh

| **Lineage** | **SARS-CoV-2 positive (%)** | **Originate** |
| --- | --- | --- |
| A | 5 (1.28) | China, India, Japan |
| A.9 | 2 (0.51) | India |
| B.1 | 29 (7.42) | UK, USA, Australia |
| B.1.1 | 76 (19.44) | UK, USA, Portugal |
| B.1.1.1 | 1 (0.26) | UK, Australia, Belgium |
| B.1.1.25 | 227 (58.06) | UK, Australia, Bangladesh |
| B.1.1.25.2 | 1 (0.26) | UK |
| B.1.1.59 | 1 (0.26) | UK, Bangladesh |
| B.1.1.60 | 1 (0.26) | UK, Austria, Bangladesh |
| B.1.148 | 1 (0.26) | UK |
| B.1.159 | 1 (0.26) | France, Bangladesh |
| B.1.2 | 2 (0.51) | USA, UK, Australia |
| B.1.36 | 30 (7.67) | India, Saudi Arabia, Bangladesh |
| B.1.5 | 1 (0.26) | UK, Spain, USA |
| B.1.5.12 | 8 (2.05) | Senegal |
| B.1.79 | 2 (0.51) | UK, India, Cyprus |
| B.1.93 | 2 (0.51) | UK, Switzerland, Spain |
| B.2.1 | 1 (0.26) | UK, Australia, USA |
| **Total** | **391** |  |

**Supplementary Table 4.** List of non-synonymous SNP present in Bangladeshi isolates

| **Non-Synonymous SNP** | **Variant** | **Annotation** | **Number of isolates acquiring mutation (%)** |
| --- | --- | --- | --- |
| A23403G | D614G | Spike | 381(97.44) |
| C14408T | P314L | RNA-dependent RNA polymerase, post-ribosomal frameshift | 365(93.35) |
| G28881A | R203K | Nucleocapsid protein | 306(78.26) |
| G28883C | G204R | Nucleocapsid protein | 306(78.26) |
| A1163T | I120F | Non-Structural protein 2 | 284(72.63) |
| G25563T | Q57H | ORF3a protein | 39(9.97) |
| C28854T | S194L | Nucleocapsid protein | 31(7.92) |
| G19723T | V35F | endoRNAse | 18(4.60) |
| C26527T | A2V | Membrane | 12(3.06) |
| C2040T | T412I | Non-Structural protein 2 | 12(3.06) |
| C25504G | Q38E | ORF3a protein | 12(3.06) |
| G4201T | M494I | Predicted phosphoesterase, papain-like proteinase | 11(2.81) |
| G11083T | L37F | Transmembrane protein | 11(2.81) |
| C2388T | T528I | Non-Structural protein 2 | 11(2.81) |
| A10323G | K90R | 3C-like proteinase | 9(2.30) |
| C2910T | T64I | Predicted phosphoesterase, papain-like proteinase | 9(2.30) |
| G8371T | Q1884H | Predicted phosphoesterase, papain-like proteinase | 8(2.04) |
| C12369T | T93I | Non-Structural Protein 8 | 8(2.04) |
| C21575T | L5F | Spike | 8(2.04) |
| A29403G | D377G | Nucleocapsid protein | 7(1.79) |
| C26895T | H125Y | Membrane | 7(1.79) |
| G21809T | V83F | Spike | 7(1.79) |
| A4503T | E595V | Predicted phosphoesterase, papain-like proteinase | 6(1.53) |
| G5950T | K1077N | Predicted phosphoesterase, papain-like proteinase | 6(1.53) |
| C21855T | S98F | Spike | 6(1.53) |
| T28144C | L84S | ORF8 protein | 5(1.27) |
| G28878A | S202N | Nucleocapsid protein | 5(1.27) |
| C21846T | T95I | Spike | 5(1.27) |
| A2870G | N51D | Predicted phosphoesterase, papain-like proteinase | 5(1.27) |
| C1593T | S263F | Non-Structural protein 2 | 5(1.27) |
| C2940T | P74L | Predicted phosphoesterase, papain-like proteinase | 5(1.27) |
| T3836A | L1191M | ORF1a polyprotein | 5(1.27) |
| C17518T | L428F | Helicase | 4(1.02) |
| G25906T | G172C | ORF3a protein | 4(1.02) |
| A10329G | D92G | 3C-like proteinase | 4(1.02) |
| G20438T | S273I | endoRNAse | 4(1.02) |
| G12170A | A27T | Non-Structural Protein 8 | 4(1.02) |
| G11222T | V84F | Transmembrane protein | 4(1.02) |
| C21707T | H49Y | Spike | 4(1.02) |
| G28960T | Q229H | Nucleocapsid protein | 4(1.02) |
| G23587T | Q675H | Spike | 4(1.02) |
| C1190T | P129S | Non-Structural protein 2 | 4(1.02) |
| G13812T | M115I | RNA-dependent RNA polymerase, post-ribosomal frameshift | 4(1.02) |
| C4113T | A465V | Predicted phosphoesterase, papain-like proteinase | 4(1.02) |
| T2211C | V649A | ORF1ab | 4(1.02) |
| C3177T | P153L | Predicted phosphoesterase, papain-like proteinase | 4(1.02) |

**GISAID acknowledgements**

We gratefully acknowledge the Authors from the Originating laboratories responsible for obtaining the specimens, as well as the Submitting laboratories where the genome data were generated and shared via GISAID, on which this research is based. All acknowledgements for the global phylogeny produced by Rob Lanfear of all GISAID submissions up to 31/7/20 can be found at <https://github.com/roblanf/sarscov2phylo/tree/31-7-20/acknowledgements>

Bangladeshi strain GISAID acknowledgements can be found in additional file Bangladesh_gisaid_acknowledgement.pdf.
